# Knowledge, Attitude, and Practice among the Healthcare Professionals regarding the myths on COVID-19 vaccination - Demystified

**DOI:** 10.1101/2021.07.30.21261378

**Authors:** Lokesh Kumar S, Zameera Naik, Arun Panwar, Sridhar M, Vaishali Keluskar, Ram Surath Kumar K

**Author notes:** **Corresponding Author:** Dr. Lokesh Kumar S, BDS., MDS., Post-graduate student, Department of Oral Medicine and Radiology, KAHER’s KLE Vishwanath Katti Institute of Dental Sciences, JNMC Campus, Belagavi- 590010, Karnataka, India., E-mail ID.

## Abstract

**Background:** COVID-19 vaccine is the mighty weapon opted by all the countries across the globe in an attempt to eradicate the fatal COVID-19 pandemic. The myths on the COVID-19 vaccine are spreading widely, causing a hindrance to this noble preventive measure. The prevalence of such myths among the healthcare professionals may be toxic and deadly.

**Aim & Objectives:** To assess the knowledge, attitude, and practice of the healthcare professionals regarding the myths on COVID-19 vaccination and to demystify them.

**Materials and Methods:** An 18-item questionnaire evaluating knowledge, attitude, and practice based on the existing myths on COVID-19 vaccination was circulated through Google Forms^®^ among the 412 healthcare professionals of six disciplines belonging to a private University. The responses obtained were subjected to statistical analysis using SPSS^®^ 20 software package.

**Results:** A total of 385 health professionals participated in this study. The majority of them had medium knowledge (165) and positive attitude (273) with the mean knowledge and attitude scores of 3.82 ± 1.55 out of 6 and 4.3 ± 1.58 out of 7 respectively. Even though 312 participants got vaccinated, 72 of them failed to receive it. The knowledge scores showed a high statistically significant difference among the participants of different designations (p=0.001), but not with gender, field, and staff with different years of experience (p>0.05). The attitude scores were statistically different among participants of fields and designation (p<0.05) but not among genders (p=0.31) and staff with different years of experience (p=0.87). Knowledge and attitude scores showed a positive linear correlation and a high statistically significant difference (p<0.001).

**Conclusion:** This study recommends more enhanced education programs on COVID-19 vaccination for the health professionals and demands an improved knowledge, attitude, and practice among them to achieve the goal of 100% vaccination so as to completely eradicate the COVID-19 pandemic.

## Introduction

The World Health Organization (WHO) was intimated by the Health Authority of China way back on 31st December 2019 about the outbreak of 27 pneumonia cases of unknown etiology in its Wuhan city of Central China.^[1]^ Since then, the viral disease had spread to the whole world involving almost all countries at an exponential rate, making it a global health emergency. WHO first termed the causative agent as 2019-novel-Corona Virus(2019-n-COV), which was later renamed by the Corona Virus Study Group to be Severe Acute Respiratory Syndrome Corona Virus-2 (SARS-CoV-2) that causes the disease called Corona Virus Disease 2019 (COVID-19).^[2]^ After thorough surveillance of the etiology, mode of spread, symptoms, severity, and fatality on a global scale, WHO declared the SARS-CoV-2/ corona virus disease (COVID-19) outbreak as a Public Health Emergency of International Concern (PHEIC) or a pandemic on 30th January, 2020.^[3]^ Human life has been affected in all dimensions, such as in terms of physical, mental, social and behavioral well-being by putting the world to halt.^[4-7]^ WHO Global COVID-19 statistics reported a total of 195,266,156 confirmed cases of COVID-19, including 4,180,161 deaths as of 28th July, 2021.^[8]^ The Governments of all the countries are taking mammoth efforts and facing a gargantuan struggle to bring the situation under control by adopting measures like lockdown, restrictions, other important political decisions, and especially mass-scale vaccination drives which seem to be a promising resort.^[9]^ Various researchers around the world jumped into the research on vaccine development since the outbreak and few reaped successes despite the isolation of 30 different strains across the globe in just 6 months.^[10]^ As of 29th July 2021, a total of 22 COVID-19 vaccines across the globe have been added within the WHO Emergency Usage Listing (EUL)/ Prequalification (PQ) process, with a few finalized among them.^[11]^ The focus of the global Target Product Profile (TPP) for COVID-19 vaccine is on vaccination for people under a high-risk category like healthcare workers, to provide long-term protection and rapid inception of immunity in outbreak settings.^[12]^ A total of 3,829,935,772 vaccine doses have been administered globally as of 27^th^ July 2021.^[8]^ India launched its vaccination drive on 16^th^ January 2021, starting with healthcare and frontline workers^[13]^, and achieved 441,912,395 vaccine doses as of 26^th^ July 2021 including the general population.^[14]^ Covishield™ from Serum Institute of India Pvt Ltd and Covaxin™ from Bharat Biotech, India were the two vaccines administered in India’s vaccination drive, where healthcare workers being vaccinated with the former, mostly. Despite the massive efforts taken by the Government to develop a safe and efficacious vaccine, hesitance to accept the vaccine among the people is still persistent.^[15]^

Myth is a folklore genre, that consists of stories/ narratives, playing an important role in people’s daily life.^[16]^ Despite the awareness campaigns among the public to get vaccinated through various platforms like social media, television, radio, newspapers, health talks by politicians, pamphlet distributions, and signboards, several myths and misconceptions are prevailing among the public regarding COVID-19 vaccines. These myths are spread by word of mouth and social media that may convince the public the other way, reducing the needed health practices resulting in dangerous health hazards. Such beliefs can be harmful to society since they cause clamor and disarray among the population. They also hamper the Government’s goals and efforts to eradicate and control the COVID-19 pandemic. Healthcare professionals play a pivotal role in creating awareness and promoting vaccination drive among the general public that demands superior knowledge about the facts on the COVID-19 vaccine. To the best of our knowledge, there are no studies in the literature that assess the prevalence of myths/misconceptions or knowledge about COVID-19 vaccination among healthcare professionals. Hence, the current study aims to assess the knowledge, attitude, and practice among healthcare professionals regarding myths/misconceptions on COVID-19 vaccination and to demystify them with facts.

## Materials and methods

Before the execution of this questionnaire study, protocols/ strategies to be followed regarding the literature search on myths/ misconceptions on COVID-19 vaccine, questionnaire design, validation, pilot study, and enrolment of study participants were clearly established by discussion among the researchers. The sample size was estimated to be 376 using the formula 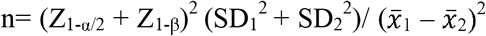 where 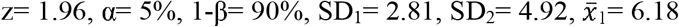, and 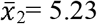 (derived from pilot study). The study was conducted among the health professionals involving the post-graduates, teaching staffs, and teaching staffs cum consultants between 20 to 65 years of age from the six constituent colleges of a private University in Belagavi District of North Karnataka, India who were included by simple random sampling, following the ethical clearance from the Institutional Research and Ethics Committee (Ref No: 1445). The sample population involved the participants from the six disciplines of healthcare namely Medicine, Dentistry, Nursing, Physiotherapy, Ayurveda and Pharmacy. A self-designed 18-item pre-validated questionnaire, based on the myths/misconceptions regarding COVID-19 vaccine known to prevail among the general public obtained from the various internet sources like the official sites of Centres for Disease Control and Prevention (Department of Health and Human Services, USA), University of Missouri Healthcare, IndiaToday etc., was used. The value of Cronbach’s alpha coefficient was 0.83. A pilot study was conducted using the prefinal version of the questionnaire among the 30 health professionals to check for internal consistency. The consistency of the questionnaire was evaluated studying the different parameters of internal consistency, test-retest reliability, and inter-rater reliability.

After getting prior permission from the respective college authorities, the questionnaire was distributed to the healthcare professionals in the Google Forms^®^ link format, which is available at https://forms.gle/CUx2pdqCuyesqFfb9, by visiting them in person and giving them enough time to fill, without any time limit. On clicking the link, this form informed the participants about the purpose, confidentiality, their voluntary involvement in the study, and also received the consent for participation (inclusion criteria) on selecting the mandatory “Yes” option to proceed with the submission. It consisted of a single page that received the participants’ demographic details of name, age, email id, gender, field, designation, years of experience if staff, informed consent, and the actual 18 questions of the study that were based on the myths and misconception on COVID-19 vaccination. All the questions were set mandatory to answer. The questionnaire comprised of 18 questions which involved 16 closed-ended questions (1 evaluated the previous experience, 6 evaluated knowledge, 7 evaluated attitude, and 2 evaluated the practice regarding COVID-19 vaccination) and 2 open-ended questions demanding to reason. The questionnaire link was sent to a total of 412 health professionals and this cross-sectional study spanned over 2 months from May 2021 to June 2021. Knowledge score was calculated based on the participants’ responses to the 6 knowledge-based questions, where the correct response was coded as “1” and the incorrect responses were coded as “0”. Hence, the maximum knowledge score was set at 6 and was graded as follows: 0-2= low, 3-4= medium, and 5-6= high. Similarly, the correct response in regards to 7 attitude-related questions were coded as “1” and the incorrect responses were coded as “0”. Thus, the maximum attitude score was set at 7 and was graded as follows: 0-3= negative and 4-7= positive. The responses from the pilot study were excluded from the main analysis.

### Statistical Analysis

The obtained data were subjected to statistical analysis using IBM-SPSS^®^ 20 software package, USA. The normality distribution of the study data was assessed by the Shapiro-Wilk test to find it out to be not normally distributed (p<0.05). Consecutively, descriptive statistics for the frequency distribution and percentage of healthcare professionals, Chi-square test for the association between the variables of the study and knowledge & attitude questions, Mann-Whitney U test for the significance among gender, and Kruskal-Wallis test for the significance among the other study variables were applied. Additionally, the correlation between the knowledge and attitude scores was evaluated by Spearman’s rank correlation coefficient test whereas, their association with the demographic details of the healthcare professionals was analyzed by simple linear regression and multivariate linear regression analysis. The statistical significance was set at p≤0.05 for all the tests.

## Results

A total of 385 responses from the healthcare professionals (185 males and 200 females) were recorded out of the 412 health professionals with a response rate of 93.44%, of which 92, 118, 44, 68, 32, and 31 participants were from fields of Medicine, Dentistry, Nursing, Physiotherapy, Pharmacy, and Ayurveda respectively. The maximum number of participants were post-graduates (304) and others being-teaching staff (23) and teaching staff cum consultants who were in clinical practice (58). Among the staff, the majority of them had an experience of 0-4 years (21) and 4-8 years (20) whereas other had above 8 years (40). The characteristics of the health professionals who participated in the study are given in Table 1. All the health professionals received the COVISHIELD™ vaccine manufactured by the Serum Institute of India Pvt Ltd. Among the 385 health professionals, 312 got themselves vaccinated and 73 did not take the vaccination. The reasons given for not getting vaccinated included the question on the safety of the vaccine, chances of testing positive even after vaccination, possible side effects & death, the emergence of different new strains of COVID-19 virus, lactating mother, and incomplete clinical trials.

**Table 1:** Characteristics of the study participants.

| **Characteristics** | **n (%)** |
| --- | --- |
| **Gender** |  |
| Male | 185 (48.1) |
| Female | 200 (51.9) |
| Total | 385 (100) |
| **Field** |  |
| Medicine | 92 (23.9) |
| Dentistry | 118 (30.6) |
| Nursing | 44 (11.4) |
| Physiotherapy | 68 (17.7) |
| Pharmacy | 32 (8.3) |
| Ayurveda | 31 (8.1) |
| **Designation** |  |
| Post-graduates | 304 (79) |
| Teaching staffs | 23 (6) |
| Teaching staff and consultants | 58 (15.1) |
| **Experience among the staffs** |  |
| 0-4 years | 21 (26) |
| 4-8 years | 20 (24.6) |
| >8 years | 40 (49.4) |
| Total | 81 (100) |

**Table 2:**
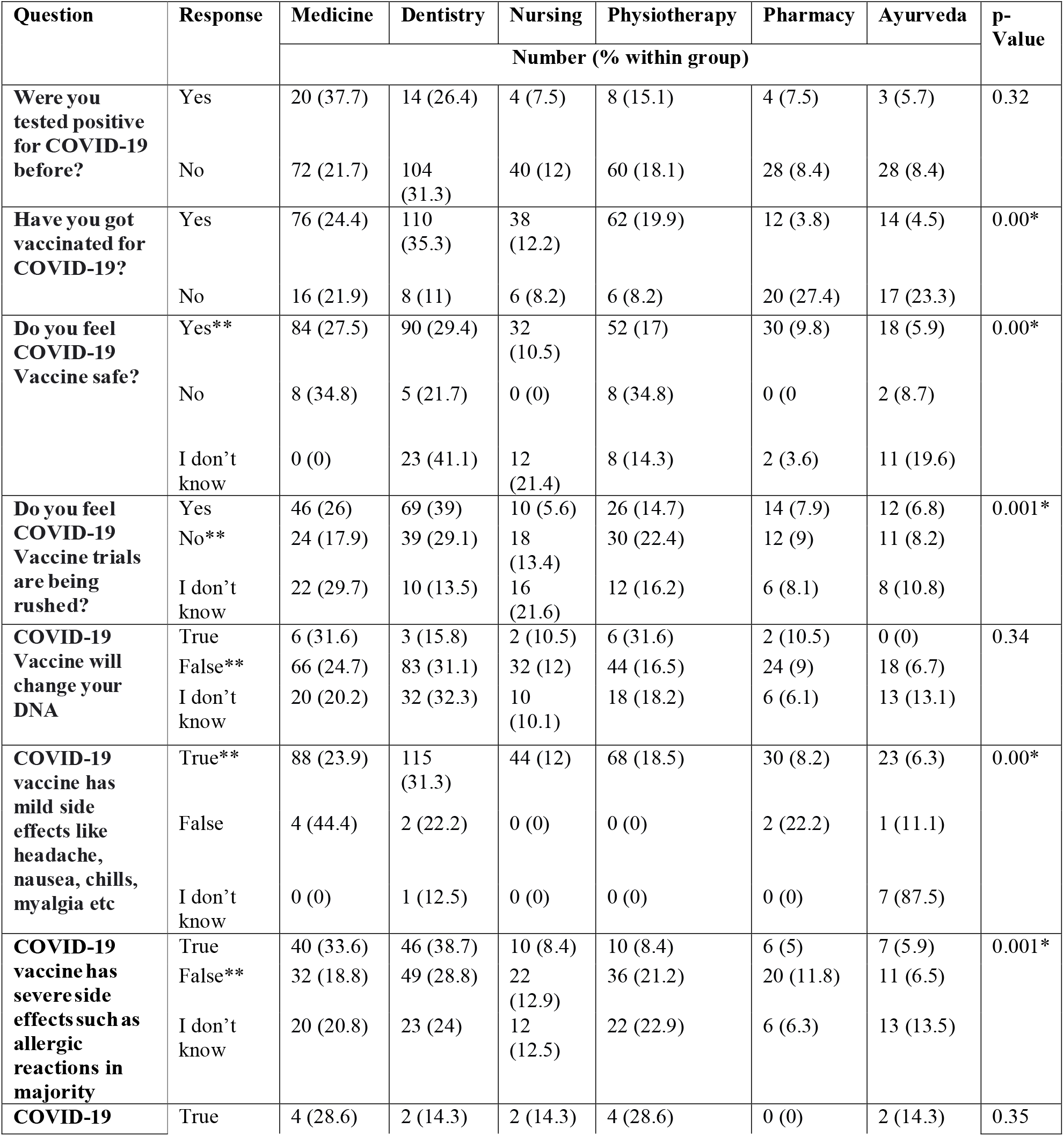
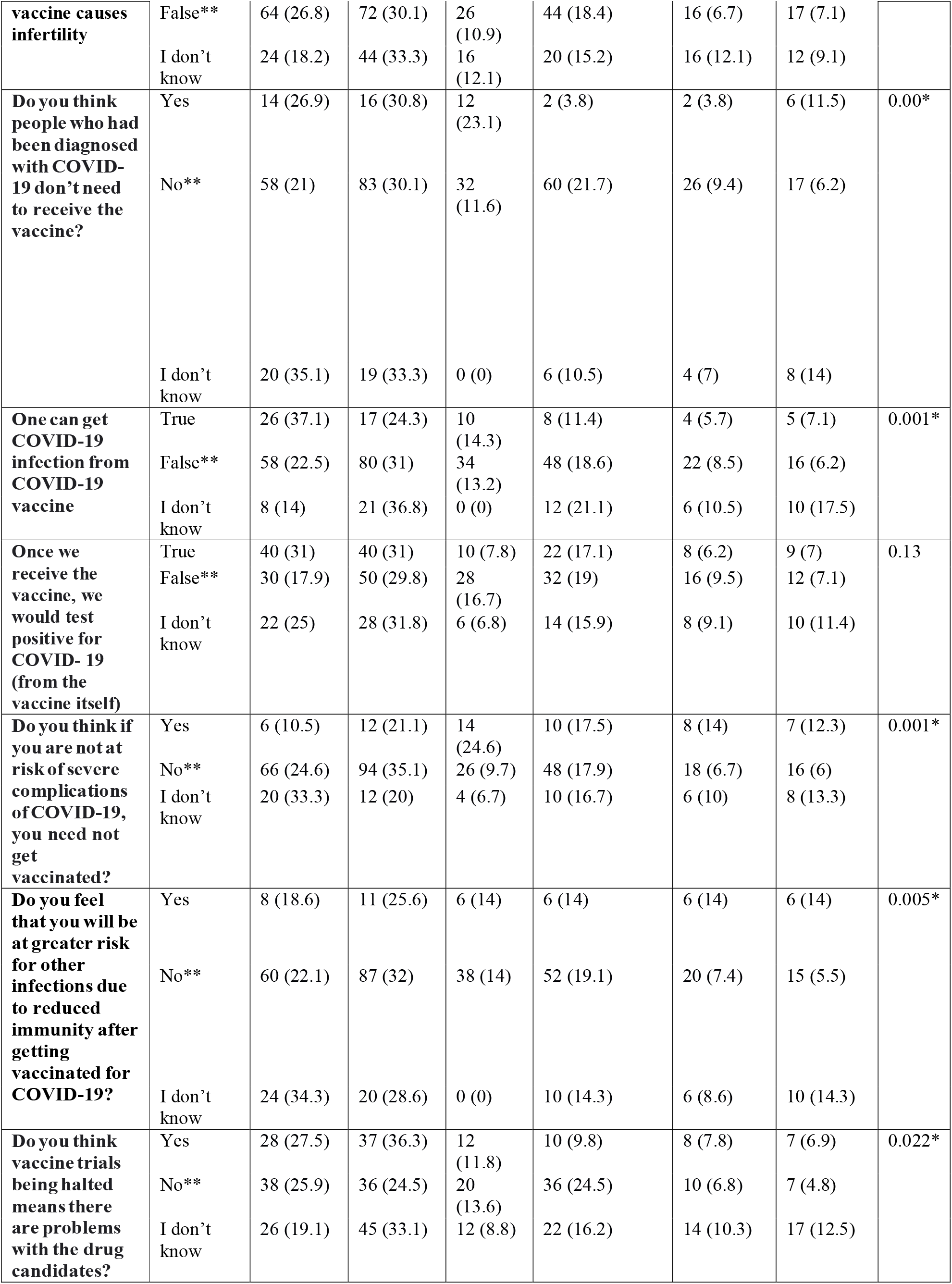
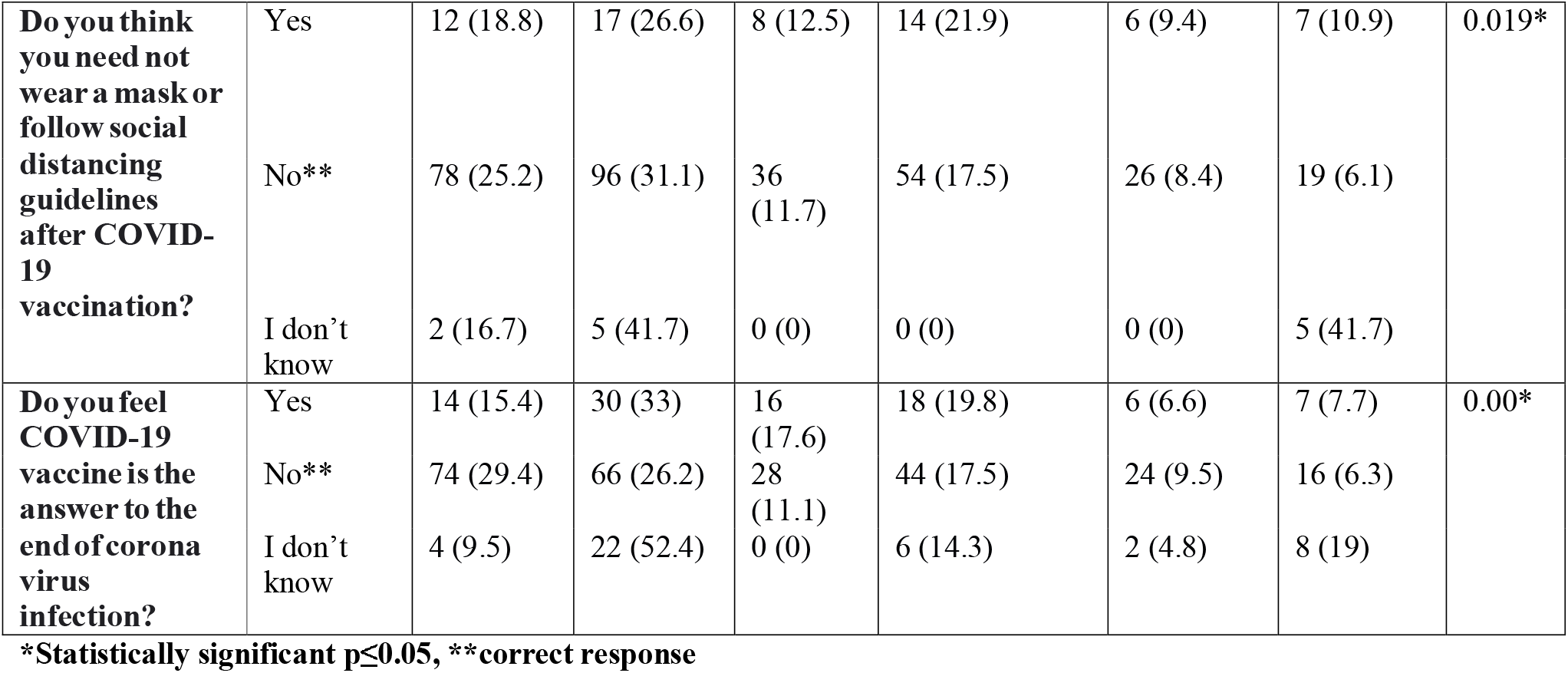
Responses for the questions on COVID-19 vaccination and its myths/ misconceptions among the health professionals.

Fifty-three of the 385 health professionals answered that they had been tested positive for COVID-19 infection before, out of which, 45 participants still took the vaccine and 8 of them did not take it. The majority of the health professionals (306) considered the COVID-19 vaccine to be safe, whereas 23 considered it to be unsafe and 56 did not know regarding the safety of the vaccine. Interestingly, 43 of the 306 participants who considered the vaccine to be safe did not get vaccinated and 14 of the 23 participants who considered the vaccine to be unsafe got vaccinated. On the other hand, 35 of the 56 participants who did not know whether the COVID-19 vaccine is safe got vaccinated. The responses of the health professionals for the questions on COVID-19 vaccination and its myths/ misconceptions are presented in Table 1.

### Knowledge about COVID-19 vaccination and its misconceptions

The majority of the health professionals (165) had a medium knowledge score whereas, 139 and 81 participants had high and low knowledge scores respectively (Chart 1). Kruskal-Wallis test depicted that there was no significant difference (p>0.05) in the knowledge scores between the health professionals of different fields, the staffs with different years of experience (0-4 years, 4-8 years, and >8years), however, showed a high statistically significant difference between the different designations (p=0.001). Mann-Whitney U test showed that there was no significant difference in the knowledge scores between the health professionals of different gender (p=0.84). (Table 3). The mean knowledge score among the health professionals was 3.82 ± 1.55 out of 6 with the highest in teaching staff cum consultant (4.64 ± 2) among designations and in staffs with the experience of 4-8 years (4.7 ± 1.34). The knowledge scores of post graduates, teaching staff, staff with experience of 0-4 years and above 8 years were 3.72 ± 1.53, 3.39 ± 1.61, 4.05 ± 1.85, and 3.95 ± 1.50 respectively.

**Chart 1:**
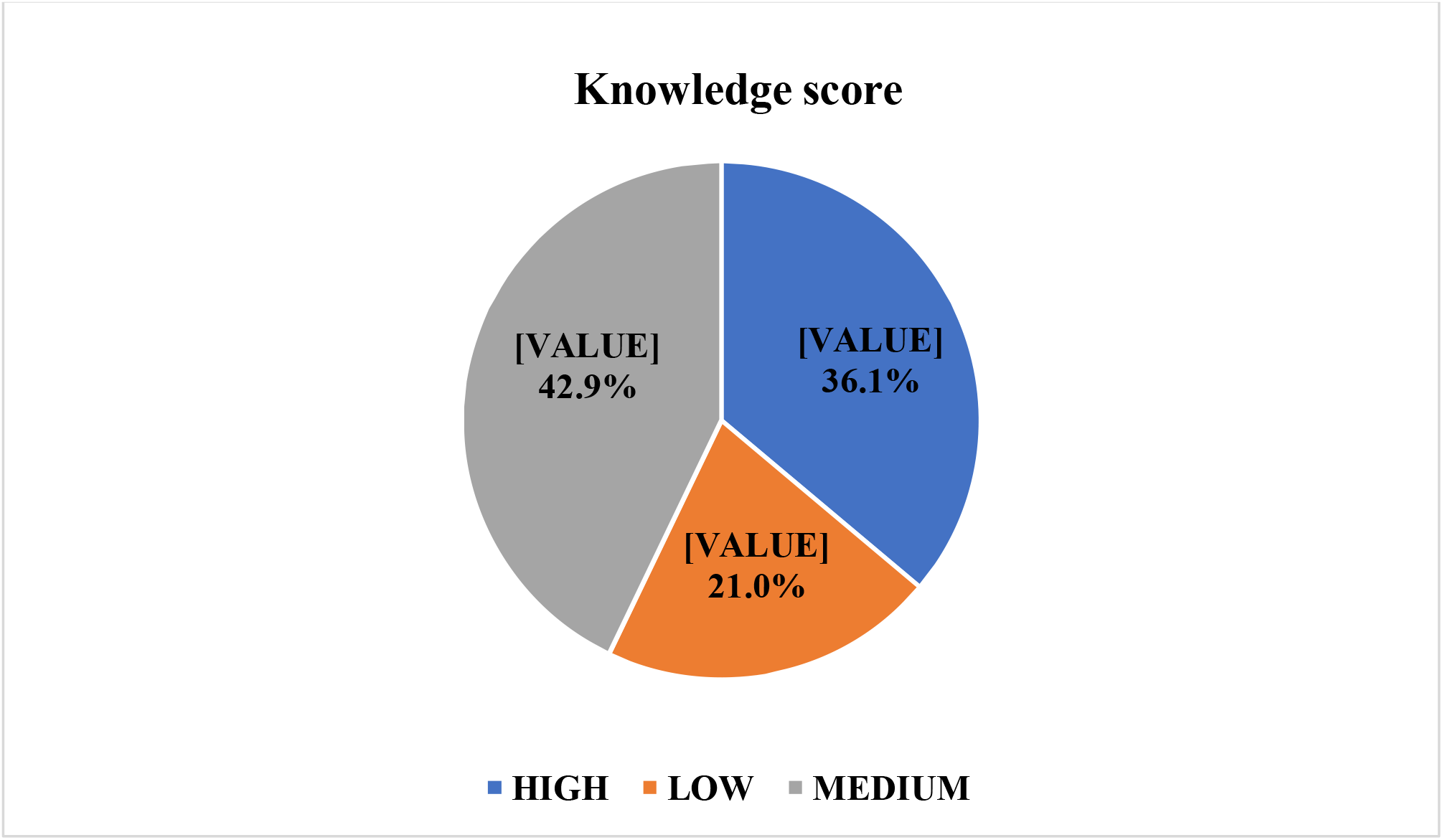
Knowledge score among the health professionals.

**Table 3:** Knowledge scores among the health professionals.

| Knowledge score | (n) mean ± SD |  |  |  |  |  |  |  |
| --- | --- | --- | --- | --- | --- | --- | --- | --- |
|  | Medicine | Dentistry | Nursing | Physiotherapy | Pharmacy | Ayurveda | Rank | p-value |
| Total** | (92) 3.67 ± 1.44 | (118) 3.89 ± 1.94 | (44) 4.23 ± 1.49 | (68) 4.00 ± 1.47 | (32) 4.00 ± 1.30 | (32) 4.00 ± 1.30 | - | 0.09 |
| Based on gender*** |  |  |  |  |  |  |  |  |
| Male | (84) 3.69 ± 1.50 | (44) 4.11 ± 2.33 | (14) 4.86 ± 1.41 | (16) 3.88 ± 1.41 | (8) 4.50 ± 0.53 | (19) 3.11 ± 1.88 | 194.18 | 0.84 |
| Female | (8) 3.50 ± 0.53 | (74) 3.76 ± 1.67 | (30) 3.93 ± 1.46 | (52) 4.04 ± 1.49 | (24) 3.83 ± 1.43 | (12) 3.17 ± 1.64 | 191.91 |  |
| Based on designation** |  |  |  |  |  |  |  |  |
| Post-graduate | (82) 3.71 ± 1.36 | (92) 3.61 ± 1.65 | (24) 4.42 ± 1.28 | (48) 3.92 ± 1.51 | (30) 4.00 ± 1.34 | (28) 2.96 ± 1.77 | 186.37 | 0.001* |
| Teaching Staffs | (2) 3.00 ± 0.00 | (13) 3.69 ± 1.49 | (2) 1.00 ± 0.00 | (6) 3.67 ± 1.86 | (0) - | (0) - | 161.15 |  |
| Teaching staff and consultant | (8) 3.50 ± 2.33 | (13) 6.08 ± 2.84 | (18) 4.33 ± 1.46 | (14) 4.43 ± 1.09 | (2) 4.00 ± 0.00 | (3) 4.67 ± 0.58 | 240.40 |  |
| Based on years of experience among staff** |  |  |  |  |  |  |  |  |
| 0-4 years | (8) 3.25 ± 2.31 | (2) 4.00 ± 1.90 | (6) 5.00 ± 1.55 | (4) 4.00 ± 1.15 | (0) - | (1) 5.00 ± 0.00 | 40.19 | 0.16 |
| 4-8 years | (2) 4.00 ± 0.00 | (6) 4.00 ± 1.90 | (6) 4.67 ± 1.03 | (6) 5.67 ± 0.52 | (0) - | (0) - | 49.25 |  |
| >8 years | (0) - | (18) 4.72 ± 1.41 | (8) 2.75 ± 1.58 | (10) 3.40 ± 1.07 | (2) 4.00 ± 0.00 | (2) 4.50 ± 0.71 | 37.30 |  |

\*Statistically significant $p \leq 0.05$ , \*\*Kruskal-Wallis Test, \*\*\*Mann-Whitney U Test.

### Attitude about COVID-19 vaccination and its misconceptions

The majority of the health professionals (273) had a positive attitude towards the COVID-19 vaccination and on the other hand, 112 participants had a negative attitude (Chart 2). There was a statistically significant difference in the attitude scores (p<0.05) between the health professionals of different fields and different designations but was not significant between the staff with different years of experience (p=0.87) when tested by the Kruskal-Wallis test. Mann-Whitney U test for the attitude scores between the male and female health professionals depicted that they were not statistically significant with a p-value of 0.31. (Table 4) The mean attitude score among the participants was 4.3 ± 1.58 out of 7 with the highest in teaching staff cum consultant among designations and in staff with experience of 4-8 years (4.80 ± 1.28). The attitude scores of post graduates, teaching staff, staff with experience of 0-4 years and above 8 years were 4.21 ± 1.61, 4.09 ± 1.47, 4.52 ± 1.40, and 4.6 ± 1.5 respectively.

**Chart 2:**
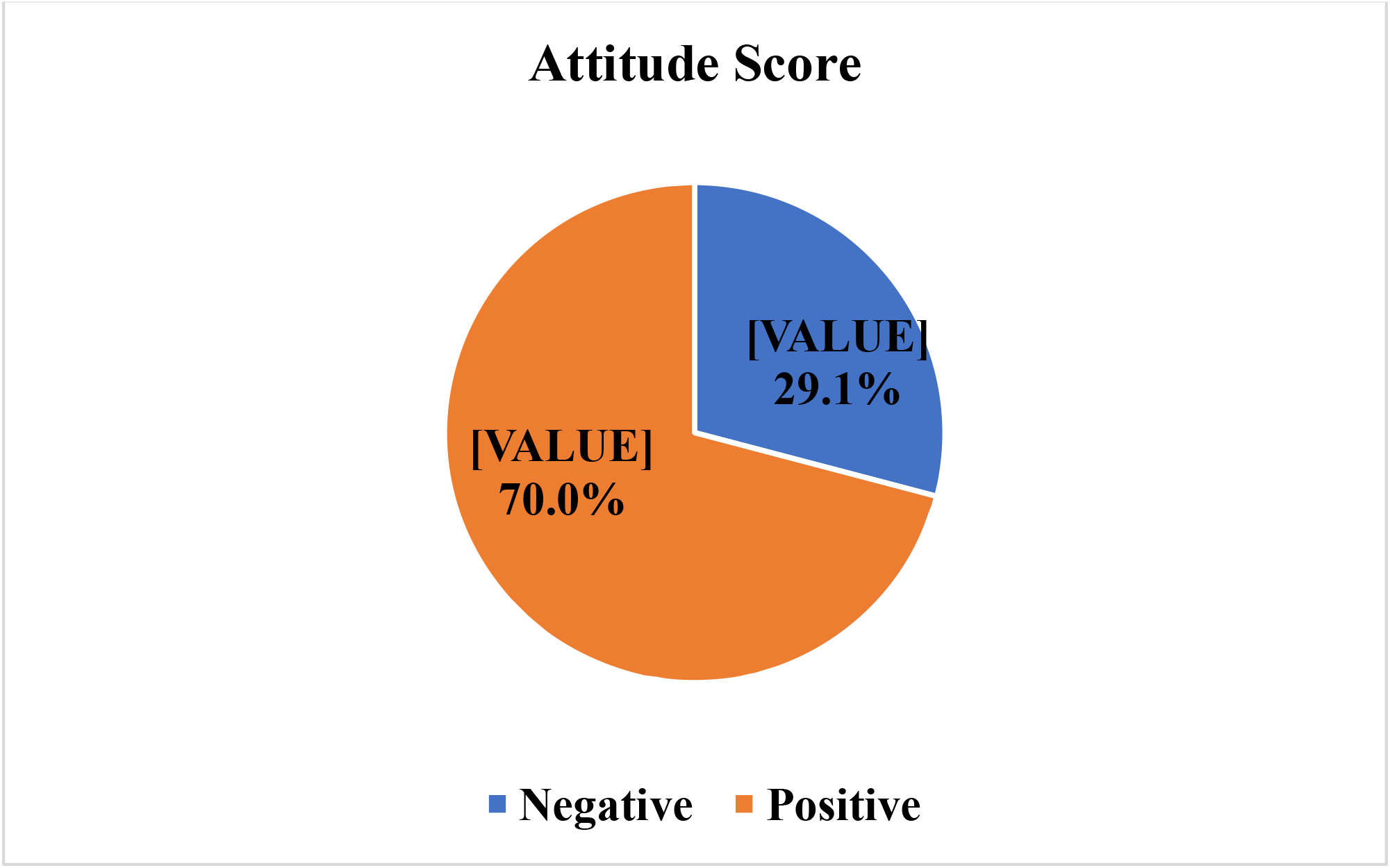
Attitude score among the health professionals.

**Table 4:** Attitude scores among the health professionals.

| Attitude score | (n) mean ± SD |  |  |  |  |  |  |  |
| --- | --- | --- | --- | --- | --- | --- | --- | --- |
|  | Medicine | Dentistry | Nursing | Physiotherapy | Pharmacy | Ayurveda | Rank | p-value |
| Total** | (92) 4.39 ± 1.33 | (118) 4.19 ± 1.55 | (44) 4.41 ± 1.45 | (68) 4.74 ± 1.85 | (32) 4.38 ± 1.13 | (31) 3.23 ± 1.84 | - | 0.002* |
| Based on gender*** |  |  |  |  |  |  |  |  |
| Male | (84) 4.52 ± 1.32 | (44) 4.11 ± 1.42 | (14) 4.14 ± 1.51 | (16) 4.63 ± 1.71 | (8) 4.25 ± 0.46 | (19) 3.11 ± 1.85 | 187.12 | 0.31 |
| Female | (8) 3.00 ± 0.00 | (74) 4.24 ± 1.64 | (30) 4.53 ± 1.43 | (52) 4.77 ± 1.91 | (24) 4.42 ± 1.28 | (12) 3.42 ± 1.88 | 198.44 |  |
| Based on designation** |  |  |  |  |  |  |  |  |
| Post-graduate | (82) 4.29 ± 1.32 | (92) 4.05 ± 1.62 | (24) 4.50 ± 1.64 | (48) 4.71 ± 1.86 | (30) 4.40 ± 1.16 | (28) 3.18 ± 1.89 | 187.40 | 0.02* |
| Teaching Staffs | (2) 6.00 ± 0.00 | (13) 4.15 ± 1.14 | (2) 3.00 ± 0.00 | (6) 3.67 ± 2.07 | (0) - | (0) - | 176.93 |  |
| Teaching staff and consultant | (8) 5.00 ± 1.31 | (13) 5.23 ± 1.01 | (18) 4.44 ± 1.20 | (14) 5.29 ± 1.64 | (2) 4.00 ± 0.00 | (3) 3.67 ± 1.53 | 228.71 |  |
| Based on years of experience among staff** |  |  |  |  |  |  |  |  |
| 0-4 years | (8) 5.25 ± 1.39 | (2) 4.50 ± 0.71 | (6) 4.00 ± 0.89 | (4) 4.50 ± 1.73 | (0) - | (1) 2.00 ± 0.00 | 56.78 | 0.87 |
| 4-8 years | (2) 5.00 ± 0.00 | (6) 4.00 ± 1.26 | (6) 5.00 ± 1.79 | (6) 5.33 ± 0.52 | (0) - | (0) - | 60.73 |  |
| >8 years | (0) - | (18) 4.94 ± 1.16 | (8) 4.00 ± 0.76 | (10) 4.60 ± 2.46 | (2) 4.00 ± 0.00 | (2) 4.50 ± 0.71 | 56.66 |  |

\*p-value ≤ 0.05, \*\*Kruskal-Wallis Test, \*\*\*Mann-Whitney U Test.

### Practices regarding COVID-19 vaccination and its misconceptions

A greater number (312, 81.04%) of the health professionals got themselves vaccinated for COVID-19 infection, but 73 (18.96%) participants did not receive the vaccine that showed a high statistically significant difference among the different fields (p<0.001) using Chi-square test. The majority of the participants (309) answered that we should still need to wear a mask and follow the social distancing guidelines after COVID-19 vaccination, on the other hand, 64 of them answered that we need not follow and 12 health professionals did not know what to be done. This was statistically significant, with a p-value of 0.019 among the health professionals of different fields.

### Association between demographic variables and knowledge/attitude scores

Simple linear regression analysis depicted a significant relationship between knowledge with designation (p=0.003, R=0.150) and years of experience among staff (p=0.026, R=0.113) but not with gender (p=0.981, R=0.001) and field (p=0.801, R=0.013). It also showed a significant relationship between attitude with designation (p=0.009, R=0.132) but not with gender (p=0.550, R=0.031), field (p=0.149, R=0.074), and years of experience among staff (p=0.067, R=0.094).

Multiple linear regression analysis revealed that the better knowledge scores were significantly associated with designation (p=0.003) but not with gender (p=0.76) and fields (p=0.68) having a positive correlation (R=0.152), whereas better attitude scores were significantly associated with designation (p=0.01, R=0.132) but not with gender (p=0.21) and field (p=0.07) having a positive correlation (R=0.166). (Table 5)

**Table 5:** Multivariate linear regression analysis for the association between demographic variables and knowledge & attitude scores of health professionals.

| **Predictor** |  | **Coefficient**<br>**$\beta$** | **SE** | **t** | **CI** | **p-value** |
| --- | --- | --- | --- | --- | --- | --- |
| **Knowledge** | **Constant** | 3.36 | 0.30 | 11.30 | 2.76 - 3.96 | 0.00* |
|  | **Gender** | 0.05 | 0.16 | 0.31 | -0.27 - 0.37 | 0.76 |
|  | **Field** | -0.02 | 0.05 | -0.41 | -0.12 - 0.08 | 0.68 |
|  | **Designation** | 0.32 | 0.11 | 2.99 | 0.11 - 0.53 | 0.003* |
| **Attitude** | **Constant** | 3.84 | 0.31 | 12.39 | 3.23 - 4.45 | 0.00* |
|  | **Gender** | 0.21 | 0.17 | 1.25 | -0.12 - 0.54 | 0.21 |
|  | **Field** | -0.10 | 0.05 | -1.83 | -0.20 - 0.01 | 0.07 |
|  | **Designation** | 0.30 | 0.11 | 2.74 | 0.08 - 0.51 | 0.01* |

**\*Statistically significant $p \leq 0.05$**

### Relationship between the knowledge and attitude scores

A positive linear correlation (r= +0.431) and a high statistically significant difference (p<0.001) between the knowledge and attitude scores among the health professionals were found by the Spearman’s rank correlation coefficient test. The field-wise correlation data are presented in table 6.

**Table 6:** **Correlation between knowledge and attitude scores among the healthcare professionals of different fields^**^.**

| **Groups** | **r** | **p- value** |
| --- | --- | --- |
| **Medicine** | +0.289 | 0.005* |
| **Dentistry** | +0.451 | <0.001* |
| **Nursing** | +0.379 | 0.011* |
| **Physiotherapy** | +0.436 | <0.001* |
| **Pharmacy** | +0.340 | 0.057 |
| **Ayurveda** | +0.640 | <0.001* |
| **Total** | +0.431 | <0.001* |

\*p-value≤0.05, \*\*Spearman's rank correlation coefficient test

## Discussion

COVID-19 vaccine is considered an efficient measure to achieve herd immunity against this fatal pandemic by various countries across the globe that have launched their vaccination drives. Despite the efforts made by the governments to get all their citizens vaccinated for COVID-19 infection, certain hurdles influence the people’s acceptance of the COVID-19 vaccine, the most important being the myths/ misconceptions that are prevalent and shared through the various social media platforms which were also faced by the previous vaccination drives of rubella, mumps, measles, and polio.^[17,18]^ In a survey conducted in 2020, almost one-third or more of the population globally answered that they might not receive the first COVID-19 vaccines questioning their efficacy, side effects, and rushing through the regulatory approval procedures.^[19]^ This study involved a questionnaire with questions, that were framed based on a few important myths/ misconceptions that are known to be widespread among the public. It was conducted among the health professional population, as they are very crucial for creating awareness and promoting the COVID-19 vaccine owing to their direct interactions with the public. The prevalence of these myths/ misconceptions on the COVID-19 vaccine among healthcare professionals may prove toxic as they are responsible for being the advocates and ambassadors for the vaccination drive.

The majority of the participants (58%) felt that the vaccine trials are being rushed (171) or they did not know regarding it (56). But the fact states that, like any other vaccine, the COVID-19 vaccine went through all the rigorous processes of safety reviews and clinical trials as prescribed by the Food and Drug Administration (FDA) agency, without skipping any of the steps although it had been developed in a short record timeframe and given a nod under EUA as biologics license approval (BLA) mandates a long time. The phase III trials were conducted on a large number of human volunteers same as any large vaccine trials in the United States and internationally.^[20]^ Vaccines generally entail a timeline of 10.7 years on an average^[20]^, and hence the COVID-19 vaccine research was built on the previous decade’s research on the corona virus than mere start after the outbreak.^[21]^ This was made possible because of the unprecedented collaboration and investment of the researchers, government, and organizations worldwide. In this current study, 267 health professionals (69.35%) answered correctly that the COVID-19 vaccine will not alter the host’s DNA. The pioneer vaccines which were granted EUA contained messenger RNA (mRNA) that instructs the cells to produce the “spike protein” that is responsible for creating the antibodies. The mRNA gets translated into polypeptides in the cytoplasm and never enters the cellular nucleus where the DNA is present. Moreover, the body disposes the mRNA after the generation of immunity and cellular DNA similar to COVID-19 virus sequences on in-vitro overexpression of LINE-1 or HIV-1 reverse transcriptase in HEK cell lines^[22]^ but, lacks evidence relevant to human clinical medicine^[21]^. About 215 (55%) health professionals answered that the COVID-19 vaccine causes severe allergic reactions such as anaphylactic reactions in the majority which is not true as it can only cause mild side effects such as fever, chills, and malaise that are known to be present with all vaccinations. The anaphylactic reactions are extremely rare, and experts recommend not to receive the vaccine if there is known history of severe allergic reactions to the ingredients of the vaccine. Around 62.07% of the participants (239) answered correctly that the COVID-19 vaccine does not cause infertility. A myth/ false information on social media stated that the vaccine makes the immune system fight against syncytin-1, a mammalian placental protein present in women which causes infertility in women.^[23]^ But, despite the spike protein and placental protein sharing a homologous amino acid sequence, it cannot produce infertility as it is short to cause such an effect. Also, there is a rumor that COVID-19 vaccines contain actual aborted fetal tissues but, such vaccines like AstraZeneca Oxford, CanSinoBio, and Johnson & Johnson have been propagated for many years, no longer contain the actual fetal tissue remnants. Even the Vatican has recommended that the health benefits of these vaccines overshadow the moral opposition of vaccines developed from these cell lines.^[24]^

Even the people who had been diagnosed with COVID-19 infection before, still need to receive the vaccine as the natural immunity may vary from person to person and it is still unknown that how long the same may last.^[25]^ The researchers are yet to understand the confounding factors completely like the longevity of the antibodies targeting the SARS-CoV-2 spike protein, which is shorter than expected actually.^[26]^ It is noteworthy that the people who had been infected and still received the vaccine had a 140-fold boost in antibodies against the spike protein from peak pre-vaccine levels.^[27]^ This was answered right by a larger number of health professionals (276/ 71.68%), and it is a fact that we can still be benefited from the vaccine despite the previous infection history. It’s a myth that one can get COVID-19 infection from the COVID-19 vaccine which was contradicted by 70 (18.1%) health professionals whereas 57 (14.1%) of them did not know. One cannot get COVID-19 infection from the vaccine as it contains the attenuated form and not the live virus.^[28]^ 129 (33.5%) of the health professionals answered that one would test positive for COVID-19 after receiving the vaccine by itself which is again a misconception. The tests for COVID-19 infection require samples from the respiratory system to detect the presence of the virus however, there is no live form of the virus in the vaccines to affect the test results. But it is possible to acquire COVID-19 infection after receiving the vaccine before the start of full protection.^[29]^

It is a misconception that the people who are not at risk for severe complications of COVID-19 infection need not get themselves vaccinated, as all need to get vaccinated regardless of the risk due to the chance of spreading to others on acquiring the disease, and this was answered right by 268 (69.6%) participants. It is thought wrong that one will be at greater risk for other illnesses after receiving the vaccine due to reduced immunity. Instead, the vaccine boosts immunity against COVID-19 infections and does not increase the risk for other diseases. None of the authorized vaccines have the live form of the virus.^[29]^ Only 147 (38.18%) of the 385 health professionals answered right that the vaccine trials being halted doesn’t mean that there are problems with the drug candidates. In a vaccine trial, all the effects including the adverse effects and even other effects that are not caused due to or related to the study to be noted and analyzed. Halting a trial and resuming it are the safety mechanisms to protect the volunteers of the trial until the effect is thoroughly investigated and studied.^[30]^ Around 80% of the participants (309) believed rightly that the practices of handwashing, masking, and following the social distancing guidelines to be followed even after the vaccination as all these measures are mandated necessary by The Ministry of Health and Family Welfare (MoHFW) until a sufficient number of populations gets vaccinated and the duration of protection of the vaccine is unknown. This highlights the importance of following the appropriate COVID-19 protocols and guidelines until further recommendations from the healthcare agencies and public health experts. Almost 23.63% of the healthcare professionals (91) answered that the COVID-19 vaccine would be an answer to the end of the corona virus which is not true. Although vaccines in the past have played a pivotal role to eradicate the small pox and reduce the incidence rate of polio, it took many years to achieve such a success. Vaccination to almost all citizens is a tedious and time-consuming process, especially with new vaccines and the virus is known to mutate. These question the long-term efficacy of the vaccine and also, the method of natural immunity cannot be resorted, as the fatality rate of COVID-19 infection is very high. Hence, believing that a vaccine would be an answer to the end of this pandemic is highly audacious at this point.

One another myth/ misconception about COVID-19 vaccination is that the vaccine has a tracking or surveillance device which was false information that was circulated in a video form on social media. A syringe maker in America, ApiJect Systems Corp^®^ has an optional variant of its product with an embedded microchip that enables the administrator to find the origin of the vaccine and however, the microchip is not injected into the body. Additionally, there is a spreading fear that vaccine trials might cause inadvertent consequences like the 1950-60s tragedy of using thalidomide for the treatment of early pregnancy nausea.^[21]^

In the current study, though 79% of the health professionals had medium to high knowledge scores, 21% of the health professionals still projected a low knowledge score level which requires additional enhancement of knowledge and awareness among the health professionals. Although 70% had a positive attitude score, 29.1% had a negative attitude score demanding a change in attitude towards the COVID-19 vaccination as they have the responsibility to eliminate the vaccine hesitancy among general public. It is also depicted that the better knowledge levels led to a better attitude towards the COVID-19 vaccination among the health professionals. It is a good sign that 81% of the health professionals had got vaccinated, in turn trusted and supported the vaccination drive.

## Limitations

This study was conducted in a smaller sample size of the health professionals of a single University. Future studies could be done, involving a larger population of health professionals from multiple centers, with a uniform sample size among the fields.

## Conclusion

Even though the participants had a high standard of knowledge, attitude and practice, there were a considerable percentage of participants with low knowledge and negative attitude. Hence, the findings of the study demand a more enhanced, stronger, and accessible health education programs on COVID-19 vaccination for the health professionals to improve their knowledge, attitude and practices regarding COVID-19 vaccination drive to reduce the vaccination hesitancy among the general public and to motivate all the citizens to get vaccinated against this deadly pandemic, thereby achieving a complete eradication in the near future.

## Data Availability

There are no data pertaining to this manuscript that are available in other online repositories.

## Acknowledgement

We would like to acknowledge the support of the respective institution heads and the study participants of KLE Academy of Higher Education and Research, Belagavi for permitting and giving their valuable time for the study respectively.

## Notes

**Conflicts of Interest:** None.

**Funding and support:** Nil.

### Competing Interest Statement

The authors have declared no competing interest.

### Clinical Trial

Not applicable as it is a cross-sectional questionnaire study

### Funding Statement

No financial support/ grant was received in any form.

### Author Declarations

Research and Ethics committee, KLE VK Institute of dental sciences, KAHER, Nehru Nagar, Belagavi, Karnataka, India. Ethical clearance approval No: 1445

## References

1. Lu H, Stratton CW, Tang Y-W. Outbreak of pneumonia of unknown etiology in Wuhan, China: The mystery and the miracle. J Med Virol. 2020;92:401-2. doi: https://doi.org/10.1002/jmv.25678

2. Gorbalenya AE, Baker SC, Baric RS, de Groot RJ, Drosten C, Gulyaeva AA, et al. Severe acute respiratory syndrome-related coronavirus: The species and its viruses –a statement of the Coronavirus Study Group. bioRxiv. 2020;2020.02.07. doi: https://doi.org/10.1101/2020.02.07.937862

3. Burki TK. Coronavirus in China. Lancet Respir. 2020;8(3):238. doi: https://doi.org/10.1016/s2213-2600(20)30056-4

4. Chopra S, Ranjan P, Singh V, Kumar S, Arora M, Hasan MS, et al. Impact of COVID-19 on lifestyle-related behavioursa cross-sectional audit of responses from nine hundred and ninety-five participants from India. Diabetes Metab Syndr. 2020;14(6):2021-30. doi: https://doi.org/10.1016/j.dsx.2020.09.034

5. Kumari A, Ranjan P, Sharma KA, Sahu A, Bharti J, Zangmo R, et al. Impact of COVID-19 on psychosocial functioning of peripartum women: a qualitative study comprising focus group discussions and in-depth interviews. Int J Gynaecol Obstet. 2021;152(3):321-7. doi: https://doi.org/10.1002/ijgo.13524

6. Mazumder A, Bandhu Kalanidhi K, Sarkar S, Ranjan P, Sahu A, Kaur T, et al. Psycho-social and behavioural impact of COVID 19 on young adults: qualitative research comprising focused group discussion and in-depth interviews. Diabetes Metab Syndr. 2021;15(1):309-12. doi: https://doi.org/10.1016/j.dsx.2020.12.039

7. Ranjan P, Kalanidhi KB, Kaur D, Sarkar S, Sahu A, et al. Psycho-social and behavioral impact of COVID-19 on middle-aged and elderly individuals: a qualitative study. J Educ Health Promot. 2021. (In press).

8. WHO Health Emergency Dashboard, WHO (COVID-19) Homepage. https://covid19.who.int/ Accessed on July 29. 2021.

9. Koirala A, Joo YJ, Khatami A, Chiu C, Britton PN. Vaccines for COVID-19: the current state of play. Paediatr Respir Rev. 2020;35:43-9. doi: https://doi.org/10.1016/j.prrv.2020.06.010

10. Yao H, Lu X, Chen Q, Xu K, Chen Y, Cheng L, et al. Patient-derived mutations impact pathogenicity of SARS-CoV-2. 2020. Available from: https://papers.ssrn.com/sol3/papers.cfm?abstract_id=3578153

11. Guidance document-Vaccines. Status of COVID-19 Vaccines within WHO EUL/PQ evaluation process.https://extranet.who.int/pqweb/sites/default/files/documents/Status_of_COVID-19_Vaccines_within_WHO_EUL-PQ_evaluation_process-16June2021_Final.pdf Accessed on July 29, 2021.

12. World Health Organization (WHO) (2020) WHO Working Group Target Product Profiles for COVID-19 Vaccines. https://www.who.int/publications/m/item/who-target-product-profiles-for-covid-19-vaccines. Accessed on July 29, 2021

13. Ministry of Health and Family Welfare. Frequently asked questions [cited 2021 Mar 20] Available from: https://www.mohfw.gov.in/covid_vaccination/vaccination/index.html Accessed on July 29, 2021

14. WHO Health Emergency Dashboard, WHO (COVID-19). Global-India. https://covid19.who.int/region/searo/country/in Accessed on July 29, 2021.

15. Harrison EA, Wu JW. Vaccine confidence in the time of COVID-19. Eur J Epidemiol. 2020;35(4):325-30. doi: https://doi.org/10.1007/s10654-020-00634-3

16. Grover S, Sahoo S, Mehra A, Avasthi A, Tripathi A, Subramanyan A, et al. Psychological impact of COVID-19 lockdown: An online survey from India. Indian J Psychiatry. 2020;62(4):354. doi: https://doi.org/10.4103/psychiatry.indianjpsychiatry_427_20

17. Vulpe SN, Rughinis C. Social amplication of risk and probable vaccine damage”: A typology of vaccination beliefs in 28 European countries. Vaccine. 2021;39(10):1508-15. doi: https://doi.org/10.1016/j.vaccine.2021.01.063

18. Ahmed SS, Schur PH, MacDonald NE, Steinman L. Narcolepsy, 2009 A (H1N1) pandemic influenza,and pandemic influenza vaccinations: what is known and unknown about the neurological disorder, the role for autoimmunity, and vaccine adjuvants. J Autoimmun. 2014;50:1-1. doi: https://doi.org/10.1016/j.jaut.2014.01.033

19. Gadoth A., Halbrook M., Martin-Blais R., Gray A., Tobin N.H., Ferbas K., et al. Assessment of COVID-19 vaccine acceptance among healthcare workers in Los Angeles. Medrxiv. 2020. doi: https://doi.org/10.1101/2020.11.18.20234468

20. Pronker ES, Weenen TC, Commandeur H, Claassen EHJHM, Osterhaus ADME. Risk in vaccine research and development quantified. PLoS One. 2013;8(3):e57755. doi: https://doi.org/10.1371/journal.pone.0057755

21. Hotez P, Batista C, Ergonul O, Figueroa JP, Gilbert S, Gursel M, et al. Correcting COVID-19 vaccine misinformation: Lancet Commission on COVID-19 vaccines and therapeutics task force members. E Clinical Medicine. 2021;33:100780.. doi: https://doi.org/10.1016/j.eclinm.2021.100780

22. Zhang L., Richard A., Khalil A., Wogram E., Ma H., Young R.A., et al. SARS-CoV-2 RNA reverse-transcribed and integrated into the human genome. Biorxiv. 2020. doi: https://doi.org/10.1101/2020.12.12.422516

23. Gong, R, Peng, X, Kang, S, Feng, H, Huang, J, Zhang, W, et al. Structural characterization of the fusion core in syncytin, envelope protein of human endogenous retrovirus family W. Biochem Biophys Res Commun. 2005;331:1193–200. doi: https://doi.org/10.1016/j.bbrc.2005.04.032

24. Vatican (2020). Congregation for the doctrine of the Faith: note on the morality of using some anti-COVID-19 vaccines https://www.vatican.va/roman_curia/congregations/cfaith/documents/rc_con_cfaith_doc_20201221_nota-vaccini-anticovid_en.html Accessed on July 29, 2021.

25. Baraniuk C. How long does covid-19 immunity last?. bmj. 2021;373. doi: http://dx.doi.org/10.1136/bmj.n1605

26. Ibarrondo FJ, Fulcher JA, Goodman-Meza D, Eliott J, Hoffmann C, Mary A, et al. Rapid decay of anti-SARS-CoV-2 antibodies in persons with mild covid-19. N Engl J Med. 2020;383(11):1085-7. doi: https://doi.org/10.1056/nejmc2025179

27. Manisty C, Otter AD, Treibel TA, McKnight A, Altmann DM, Brooks T, et al. Antibody response to first BNT162b2 dose in previously SARS-CoV-2-infected individuals. Lancet. 2021;397(10279):1057-8. doi: https://doi.org/10.1016/s0140-6736(21)00501-8

28. World Health Organization, EPI. WiN, Infodemic management. Update on COVID-19 vaccines & immune response dated 28^th^ February 2020.https://www.who.int/docs/default-source/coronaviruse/risk-comms-updates/update52_vaccines.pdf?sfvrsn=b11be994_4 Accessed on July 29, 2021.

29. Centres for Disease Control and Preventions. Myths and facts about COVID-19 Vaccines. https://www.cdc.gov/coronavirus/2019-ncov/vaccines/facts.html Accessed on July 29, 2021.

30. Dr. Pavithra Venkatagopalan. Debunking the myth related to COVID-19 vaccines dated February 4, 2021. https://www.indiatoday.in/information/story/debunking-myths-related-to-covid-19-vaccines-1763959-2021-01-29 Accessed on July 29, 2021.

